# Nanopore sequencing panel for saliva-based host pharmacogenomic screening in anti-tubercular therapy

**DOI:** 10.64898/2026.09.02.26362034

**Authors:** Priyanka Yadav, Swarup A. V. Shah, Aishwarya S. Babu, Minal Paradkar, Shruthi Vasanthaiah, Karthick Vasudevan, Prerna R. Arora, Rohan V. Lokhande, Heeral U.B. Pandya, Paolo Denti, Camilla Rodrigues, Jason R. Andrews, Akhilesh Pandey, Jeffrey A Tornheim, Tester F. Ashavaid, Renu Verma, MDR-TB MUKT and RePORT India Study Teams

**Author notes:** Corresponding Author(s): Renu Verma, PhD, Faculty Scientist, Institute of Bioinformatics, Bangalore, Karnataka, India, 560066, Tester Ashavaid, PhD, Consultant Biochemist, PD Hinduja Hospital and Medical Research Centre, Mumbai, Maharashtra, India, 400016, Jeffrey A Tornheim, MD, MPH, Associate Professor, Johns Hopkins University School of Medicine, Baltimore, Maryland, USA, MD 21205. Shared contributions: P.Y and S.S contributed equally to this work.

## Abstract

**Rationale:** Host genotypes can predict subtherapeutic anti-tubercular drug exposures and treatment-associated toxicities. Screening for these variants could enable personalized dosing, but scalable assays for second-line drugs are lacking.

**Objectives:** We developed a nanopore sequencing panel to detect host variants affecting anti-tuberculosis drug troughs and toxicities, and evaluated its performance as a saliva-based screening tool.

**Methods:** We designed a 16-plex panel targeting 23 variants (21 clinically validated, 2 predicted actionable) relevant to linezolid, bedaquiline, clofazimine, moxifloxacin, and ethambutol exposure. We first sequenced 50 Coriell DNA (1000 Genomes Project) to benchmark accuracy against Illumina, then sequenced saliva from 202 individuals treated for drug-resistant tuberculosis in India using MinION Mk1C (R10.4). Plasma trough concentrations and toxicity frequencies were stratified by genotype. Data were analyzed using in-house pipelines.

**Measurements and Main Results:** The panel showed high coverage in saliva (median 3,609X). Several suggestive genotype-phenotype trends reached nominal significance in distinct subsets. Among patients on high-dose moxifloxacin (800mg daily), UGT1A1 rs3755319 A>C was associated with higher troughs in heterozygotes (6/14, p<0.01) and homozygous alternates (4/14, p<0.05). Among patients with linezolid-associated toxicity dose-reduced to 300mg, ABCB1 rs2032582 A>C homozygous alternates (7/98) had significantly lower Cmin versus wild-type (p<0.05) and heterozygotes (p<0.01); neither association held at standard dosing. Linezolid toxicity was more frequent among ABCB1 rs1128503 A>G heterozygotes versus homozygous reference (58.3% vs. 29.1%), and UGT1A1 rs4148323 G>A heterozygotes showed higher moxifloxacin toxicity rates than wild-type (42.9% vs. 14.3%).

**Conclusions:** Portable, saliva-based sequencing reliably detects pharmacogenetic variants and could inform pre-treatment screening for drug exposure or toxicity.

**At a glance summary:** *Scientific knowledge on the subject:* Interindividual variability in anti-tuberculosis plasma drug concentrations may contribute to poor outcomes, relapse, and toxicity, particularly with second-line regimens. Variants in host pharmacogenes partly explain subtherapeutic exposure and adverse effects and can be leveraged to identify individuals at risk, enabling dose optimization. However, pharmacogenes for second-line drugs remain underexplored, often assessed using limited variants in small populations despite substantial ethnic diversity. Scalable assays evaluating multiple variants are needed to enable routine screening for pharmacogenomics-guided treatment in clinical practice.

*What this study adds to the field:* We developed a custom Nanopore sequencing panel targeting pharmacogenes for five second-line anti-tuberculosis drugs, incorporating both validated and predicted pharmacogenomic variants. The panel showed 100% concordance with Illumina whole-genome data (n=50) and was clinically validated on saliva samples from 202 patients receiving guideline-concordant, susceptibility-guided multidrug therapy for rifampin-resistant tuberculosis. The panel generated high-quality data from low DNA input, supporting scalable and noninvasive screening. This allowed for 174 distinct assessments of the impact of genotype on drug concentration and 152 assessments of association between genotype and clinical toxicity, with 3 associations demonstrating significance in subpopulations of people treated for MDR-TB. Specifically, *UGT1A1* and *ABCB1* polymorphisms were associated with moxifloxacin and linezolid trough concentrations at the extremes of the doses prescribed (800mg and 300mg daily, respectively). Carriers of an *ABCB1* variant suggested a higher frequency of high-grade linezolid-associated toxicity. Overall, the panel provides baseline evidence for further validation of these markers in large-scale pharmacokinetic studies, as well as first clinical data on predicted variants that warrant screening in larger cohorts, demonstrating suitability for use on low-cost, portable Nanopore sequencers with shorter turnaround times.

## INTRODUCTION

Tuberculosis remains a leading cause of morbidity and mortality worldwide. According to recent WHO data, treatment success rates are 88% for drug-sensitive TB and 68% for rifampin-resistant or multidrug-resistant tuberculosis (MDR-TB) (1,2). Most antituberculosis drugs are prescribed using standardized flat or weight-banded doses, and dose reductions for agents such as linezolid, bedaquiline, moxifloxacin, and clofazimine are generally reserved for profoundly underweight patients or patients with toxicities. However, pharmacokinetic (PK) studies demonstrate wide interindividual variability in plasma drug concentrations with standard dosing (3–6). Variable exposures with standard dosing can lead to suboptimal levels of first-line tuberculosis drugs, which have been associated with poor outcomes, including treatment failure and relapses (7,8).

Variable plasma drug levels of anti-tubercular drugs have been reported by several studies (7–13). In a cohort of 404 patients on first-line therapy, 85% had subtherapeutic 2-hour concentrations, among which 19% experienced poor outcomes (7). In a South African cohort, individuals with at least one subtherapeutic plasma drug concentration had a 14-fold higher risk of microbiological failure, death, or relapse (10). Variability in drug exposure appears to be even more critical in second-line regimens, which have greater pharmacokinetic variability, longer duration, and fewer options for substitution (11–13). A pediatric MDR-TB study from Pakistan also demonstrated wide variability in drug plasma concentrations, with only 38% of 24 children on levofloxacin achieving therapeutic concentrations (13).

Adverse drug reactions (ADRs) to anti-tuberculosis drugs also significantly contribute to poor treatment outcomes, including increased healthcare costs (14). ADRs may lead to non-adherence or treatment discontinuation, thereby raising risks of failure, relapse, and resistance (15). Among patients on first-line drugs, between 8-30% experience at least one ADR (16). Liver enzyme elevations and drug-induced liver injury are the most frequent adverse reactions, occurring in up to 30% of patients receiving standard therapy (17,18). The frequency is much higher with second-line regimens for drug-resistant TB (40-96%) (19,20). In a study involving 98 patients on second-line therapy, 119 adverse events were recorded, affecting 46.9% (46/98) of participants (20).

Variants in drug-metabolizing enzymes and transporters may alter protein function and expression, resulting in interindividual differences in exposure and adverse reactions (21). For linezolid, *Cytochrome P450 family 3 subfamily A member 5* (*CYP3A5)* polymorphisms have been linked to altered drug disposition, potentially increasing the risk of linezolid underexposure in affected patients (22). Genetic variants in the *UDP glucuronosyltransferase family 1 member A gene complex* (*UGT1A1)* gene are shown to reduce moxifloxacin clearance and may increase the risk of drug-induced liver injury (5). Variants in the *ATP binding cassette subfamily B member 1* (*ABCB1)* gene can modify P-glycoprotein function and thereby influence the absorption and disposition of linezolid and moxifloxacin (5, 23).

Pre-treatment screening for patients at risk of subtherapeutic exposure or adverse reactions could improve outcomes. Therapeutic drug monitoring, the current standard, requires repeated blood sampling and often expensive HPLC/mass spectrometry platforms with cold-chain requirements, limiting feasibility in high-burden, resource-limited settings. Immunoassays exist only for linezolid and amikacin, leaving most anti-tuberculosis drugs without monitoring options. A pharmacogenomic (PGx) screening tool offers a scalable, cost-effective alternative. While similar benefit remains undemonstrated for MDR-TB, PGx-guided isoniazid dosing has shown benefit in a randomized trial, reducing hepatotoxicity in slow acetylators from 78% to 0% and treatment failure in rapid acetylators from 38% to 15% (24).

Most TB pharmacogenomic studies focus on first-line drugs, with limited data for second-line agents, which carry greater pharmacokinetic variability and toxicity risk. We developed a targeted Nanopore sequencing (tNGS) panel to screen pharmacogenes for second-line anti-tubercular drugs using saliva for noninvasive, pre-treatment testing. Alongside established pharmacogenomic variants, the panel includes markers predicted to affect drug metabolism or toxicity, broadening utility given scarce second-line pharmacogenomic data. The panel was validated in MDR-TB cohort participants by comparing genotypes from frozen saliva with plasma drug concentrations and observed toxicities, and performance was assessed on both SpotON and Flongle flow cells to enhance scalability and turnaround time.

## METHODS

### Study cohort and ethical approval

Clinical samples analyzed in this study were collected from participants in a single-site observational cohort of adults and adolescents aged ≥15 years recruited from the outpatient chest clinic of PD Hinduja Hospital and Medical Research Centre, Mumbai, India, at the beginning of susceptibility-guided MDR-TB treatment between 2017 and 2022. Participants received 18-24-month regimens selected according to contemporaneous Indian national guidelines (25) with adjustments according to susceptibility profiles obtained at the start of treatment. Ethical approval was obtained from the ethics committee of PD Hinduja Hospital, Mumbai (IRB00012235), Johns Hopkins University (FWA00005752), and the Institute of Bioinformatics, Bangalore (EC/NEW/INST/2021/2463). All participants aged 18 years or older provided written informed consent at enrolment. Participants aged 15-17 years provided assent for study participation, with consent provided by their legal guardians. Following their 18^th^ birthdays, all participants still in the study were reconsented as adults. Detailed clinical features and treatment history of the study participants are summarized in **Table 1**.

**Table 1:** Clinical characteristics of TB patients from MUKT cohort.

| **Participant Characteristic** | **n (%)** |
| --- | --- |
| Age, Median (IQR) | 27 (21.0-35.8) |
| Female Sex | 131 (64.9) |
| Pulmonary TB (Any) | 130 (64.3) |
| Extrapulmonary TB (Any) | 89 (44.1) |
| BMI <18.5 | 74 (36.6) |
| Diabetic | 33 (16.3) |
| HIV Positive | 5 (2.5) |
| Cavitary Lung Disease | 84 (41.6) |
| Alcohol Use | 15 (7.4) |
| Tobacco Use | 6 (3.0) |
| Prior TB | 52 (25.7) |
| Fluoroquinolone Resistant TB | 121 (59.9) |
| Injectable Resistant TB | 20 (9.9) |
| Linezolid Resistant TB | 8 (4.0) |
| Ever Prescribed Linezolid | 187 (92.6) |
| Ever Prescribed Moxifloxacin | 177 (87.6) |
| Ever Prescribed Bedaquiline | 92 (45.5) |
| Ever Prescribed Clofazimine | 187 (92.6) |
| Ever Prescribed Ethambutol | 85 (42.1) |
| Good Treatment Outcome | 177 (87.6) |
| All-Cause Mortality | 10 (5.0) |
| TB-Related Death | 8 (4.0) |
| Any Grade 3+ Bedaquiline-Associated Toxicity | 13 (14.1) |
| Any Grade 3+ Linezolid-Associated Toxicity | 58 (31.0) |
| Any Grade 3+ Moxifloxacin-Associated Toxicity | 27 (15.3) |
| Cytopenias (Any) | 153 (81.8) |
| Cytopenias (Grade 3+) | 18 (9.6) |
| Hyperpigmentation | 103 (55.1) |
| Joint Pain (Any) | 68 (38.4) |
| Joint Pain (Grade 3+) | 2 (1.1) |
| Liver Injury (Any) | 15 (7.4) |
| Liver Injury (Grade 3+) | 9 (4.5) |
| Neuropathy (Any) | 71 (38.0) |
| QTc Prolongation (Grade 3+) | 29 (14.4) |
| Vision Loss (Grade 3+) | 2 (1.1) |

MDR-TB regimens and dosing decisions were made by treating clinicians independent of study participation based on drug resistance profile and clinical comorbidities. Ethambutol was dosed according to weight bands, following Indian National guidelines (25). Bedaquiline treatment included a 14-day loading dose of 400 mg daily followed by 200 mg thrice-weekly on Mondays, Wednesdays, and Fridays for a total 6-month bedaquiline course. Linezolid was started at a dose of 600 mg daily and reduced to 300 mg daily for participants who developed linezolid-associated toxicity (peripheral neuropathy or cytopenias) and found symptoms manageable at the lower dose, while it was discontinued in the setting of either severe neuropathy or optic neuritis. Moxifloxacin was mostly provided at a daily dose of 400 mg, but was intensified to either 600 or 800 mg at clinician discretion. Clofazimine was prescribed at a dose of 100mg daily for the duration of therapy (25).

### Toxicity Assessment

Scheduled clinical evaluations were performed within 1 week of treatment initiation, two weeks after enrolment, at monthly intervals for the first six months, then quarterly until the end of therapy. At each evaluation, participants self-reported neuropathic symptoms (e.g., pain, tingling, numbness) and changes in skin color. Clinicians tested 5g monofilament and 128Hz vibration testing of both feet, visual acuity by Snellen and Ishihara plates at each visit. Transaminase levels, complete blood cell counts, and ECGs were evaluated regularly, with the corrected QT interval measured by Fridericia criteria. Each toxicity was classified as “occurred at any time during therapy” and as “occurring at grade 3 or higher at any time during therapy” following DAIDS categorization (26) **(Details in online supplement).**

### Plasma drug concentration testing

Participants provided blood for drug concentration testing at 1, 2, 6, and 12 months during treatment. At each visit, pre-dose samples (troughs) were collected prior to observed daily doses. The samples were immediately placed on ice, and plasma was separated by centrifugation within 30 minutes before storage at -80°C until measurement. Linezolid assessments were performed using a commercially available immunoassay (ARK™ Linezolid Assay) according to the manufacturer’s instructions, and moxifloxacin, bedaquiline, and clofazimine were evaluated by tandem mass spectrometry. All plasma concentration values below the limit of detection (LOD) for each drug were imputed as LOD/2 before non-parametric statistical testing described below. All testing was performed at the P.D. Hinduja Hospital and Medical Research Center in Mumbai, India (27–29).

### Saliva sample collection and DNA isolation

Before saliva collection, participants were asked not to eat, drink, or smoke. Saliva donors rinsed their mouths with water before collection of approximately 5mL of saliva in a sterile container. Saliva was stored at -80 °C until DNA extraction using the QIAamp DNA Blood Mini Kit (Cat. no. 51104) according to the manufacturer’s instructions (**Details in online supplement**). Approximately 50 ng of purified DNA was used for library preparation.

### Selection of pharmacogenes

We reviewed published literature and pharmacogenomic databases (30, 31) to identify variants affecting drug metabolism or toxicity of anti-tubercular drugs. The panel included known and predicted functional variants in drug-metabolizing enzymes that may alter plasma concentrations or ADRs. In total, we selected 23 SNPs across 12 pharmacogenes. The prevalence of pharmacogenic variants is presented in **Figure 1B (Figure 1A, Supplementary Table 1).**

**Figure 1:**
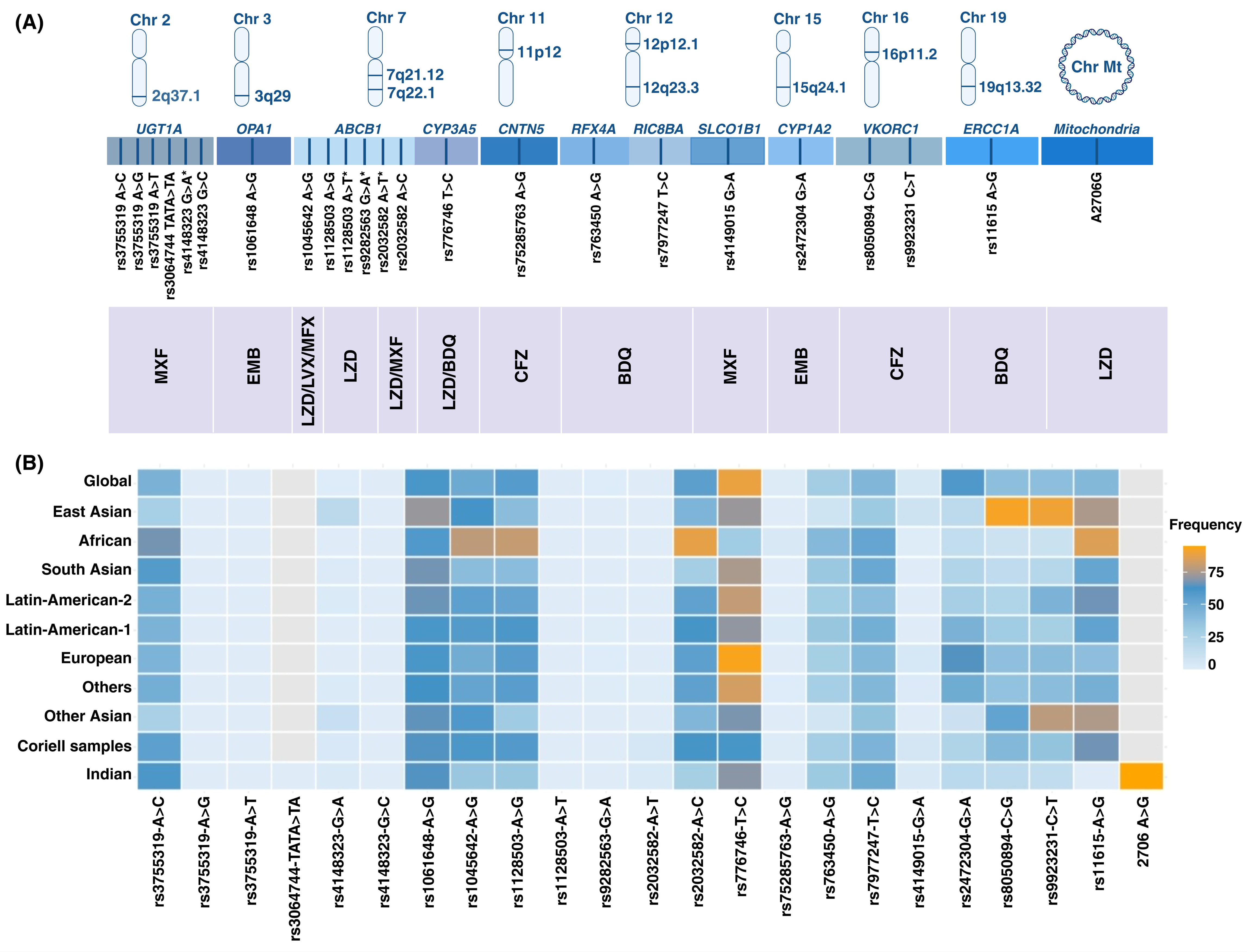
(A) Nanopore sequencing based pharmacogenomics panel for anti-tuberculosis (TB) drugs. The panel shows genes with targeted single-nucleotide polymorphisms (SNPs; n=23) along with their chromosomal positions and reference SNP cluster IDs (rsIDs). Anti- TB drugs linked to these pharmacogenomic variants are listed in the bottom row. Chr- Chromosome; Mt- Mitochondria; MXF- Moxifloxacin; EMB- Ethambutol; LZD- Linezolid; LVX- Levofloxacin; BDQ- Bedaquiline and CFZ- Clofazimine. (B) Global distribution of mutation frequency across different populations in the genes associated with the Moxifloxacin, Ethambutol, Linezolid, Levofloxacin, Bedaquiline, and Clofazimine. X-axis lists rsIDs associated with pharmacokinetic variation and toxicity, and the Y-axis indicates minor allele frequency as per the Allele Frequency Aggregator database in different populations.

### Nanopore sequencing panel design

We designed primers to amplify regions of 16 genomic targets. Both forward and reverse primers were attached to nanopore-compatible 5′ and 3′ anchor sequences suitable for the amplicon sequencing library preparation workflow. Amplicons ranging from 445 to 826 base pairs were designed using Beacon Designer software (Premier Biosoft International, version 8). Details of the anchored primer sequences are provided in **Supplementary Table 2**. The final 16-plex PCR was performed in a 50μL reaction containing LongAmp Taq DNA polymerase (NEB) and 2.5M betaine (Sigma), with optimized primer concentrations in the 0.20-0.22μM range. Cycling conditions were 94°C for 2min; 30 cycles of 94°C for 30sec, 58°C for 60sec, and 65°C for 90sec; followed by a final extension at 65°C for 10 minutes. Amplicons were purified using AMPure beads and eluted in 50μL nuclease-free water. The purified products were quantified using Qubit prior to downstream library preparation.

### DNA samples for panel development and analytical validation

For development and analytical validation of the Nanopore pharmacogenomic panel, we used commercially available Coriell DNA samples (n=50) from the 1,000 Genomes Project. Illumina whole-genome sequencing data for these samples were used as a reference to assess the variant-calling accuracy of the Nanopore panel relative to Illumina sequencing. To determine the limit of detection, a single Coriell DNA sample was sequenced at five input amounts (100, 50, 25, 12.5, and 6.25ng/μL), with all dilutions run in technical replicates.

### MinION library preparation and sequencing

The library was prepared using the SQK-LSK114 Ligation Sequencing Kit (Oxford Nanopore Technologies). Briefly, the PCR purified samples were barcoded using a Nanopore PCR barcoding expansion (EXP-PBC096 PCR Barcoding Expansion) followed by DNA repair and end-prep using NEB Next FFPE DNA Repair Mix and NEB Next Ultra II End repair/dA-tailing module reagents in accordance with the manufacturer’s instructions. Adaptor ligation was performed using Adapter Mix F (AMX-F) and Quick T4 Ligase. The samples were sequenced on a MinION Mk1C sequencer (**Details in the online supplement**).

### Global mutation frequency analysis

We performed a global data search to understand mutation frequency across different regions worldwide. Mutation frequencies were accessed from the ALFA (Allele Frequency Aggregator) database from ClinPGx (30). The mutation frequencies were analyzed to assess the prevalence of these variants in various populations and to determine whether the prevalence varies across populations.

### Sequencing data analysis

De-multiplexing and real-time basecalling were performed using the in-built MinKNOW software (Version 25.05.14) with the onboard basecalling software Guppy (Version 6.5.7). The run was configured for high-accuracy basecalling (cutoff >9). Mapping was performed by aligning the reads to a multi-FASTA file containing the concatenated sequences of the amplicons included in the panel. For coverage analysis, the reference FASTA sequence included both the forward and reverse strands of all amplicons used. FASTQ files containing passed reads were compressed and uploaded to an in-house pipeline for coverage calculation.

### Variant calling and haplotype phasing

Oxford Nanopore FASTQ reads from genomes were aligned to the human reference genome (GRCh38) using minimap2 with ONT-optimized parameters. In order to ensure high levels of accuracy, SAM files were converted to BAM format and filtered using a minimum MAPQ factor of 60, and sorted and indexed using samtools. For each genome, unphased VCF files were generated using Clair3 (32), a deep learning-based variant caller optimized for long-read sequencing, executed with ONT-specific models and multi-threaded processing. The resulting unphased variants were then subjected to haplotype phasing using WhatsHap (33), which resolves heterozygous variants into phased haplotypes using read-level information. The final output consisted of phased VCF files and a consolidated variant dataset, generated by merging variant calls across all genomes using a custom processing script. In addition, a manual variant database containing chromosomal positions and allelic information was constructed and used to extract and annotate variant information from the phased VCF files by matching against the dbSNP138 VCF reference for concordance analysis (31). These datasets were subsequently used for downstream population-level analysis.

### Statistical Analyses

To correlate drug concentrations by pharmacogenomic variants, the presence of each variant was classified as homozygous reference, heterozygous, or homozygous alternate, then correlated with drug concentrations using the Kruskal-Wallis test of statistical difference. Pre-dose drug concentrations obtained after 1-2 months of MDR-TB treatment were compared between genotypes. In a sensitivity analysis, the same comparisons were repeated for all drug concentrations measured throughout MDR-TB treatment (months 1, 2, 6, and 12). In addition, multiple doses of two study drugs (linezolid and moxifloxacin) were prescribed by clinicians. In addition to analyzing linezolid and moxifloxacin drug concentrations among participants prescribed each specific dose (300 or 600 mg for linezolid and 400, 600, or 800 mg for moxifloxacin), differences by genotype were re-analyzed after normalizing concentration results to a “standard” dose of 600mg daily for both drugs to evaluate differences by genotype across the dose range. Both the Kruskal-Wallis and the Cochran-Armitage Trend tests were used to assess the associations and trends, respectively, between genotype groups and each toxicity reported. Due to small sample sizes, toxicity assessments for moxifloxacin and linezolid were not stratified by dose of drug prescribed. For these analyses, we tested only preselected pharmacogenes for the corresponding drug. Differences were considered statistically significant at p<0.05, and due to the large number of parallel analyses with relatively small sample sizes were not corrected for multiple comparisons. Drug concentrations and toxicity analyses were performed using R Statistical Software (v4.5.0, R Core Team, 2025).

## RESULTS

### Nanopore PGx panel performance and concordance with Illumina

All amplicons in the 16-plex PGx panel achieved coverage depths greater than 50X at all input DNA quantities (100, 50, 25, 12.5, and 6.25ng/μL) of Coriell genomic DNA. Median coverages across this dilution series were 2,477X, 965X, 1,550X, 2,002X, and 1,967X, respectively. To assess performance on lower-throughput hardware, the same dilutions were sequenced on a Flongle flow cell and compared with a standard SpotON flow cell. The Flongle configuration (77 pores, 24-hour run, 300 MB) yielded a median coverage of 1,356X (interquartile range, IQR: 423-2,251), whereas the SpotON configuration (1,481 pores, 48-hour run, 4 GB) yielded 1,523X (IQR: 410-5,533). (**Supplementary Figure 1**)

We next sequenced 50 Coriell DNA samples from the 1000 Genomes Project using the 16-plex PGx panel and benchmarked variant-calling performance against Illumina whole-genome sequencing. The median sequencing depth across the 16 amplicons was 1,921X (IQR: 511-5,577), with 94.7% of amplicons covered at >100X and 76% at >500X (**Figure 2**). Variants identified with the Nanopore PGx panel showed 100% concordance with those obtained from Illumina whole-genome sequencing.

**Figure 2:**
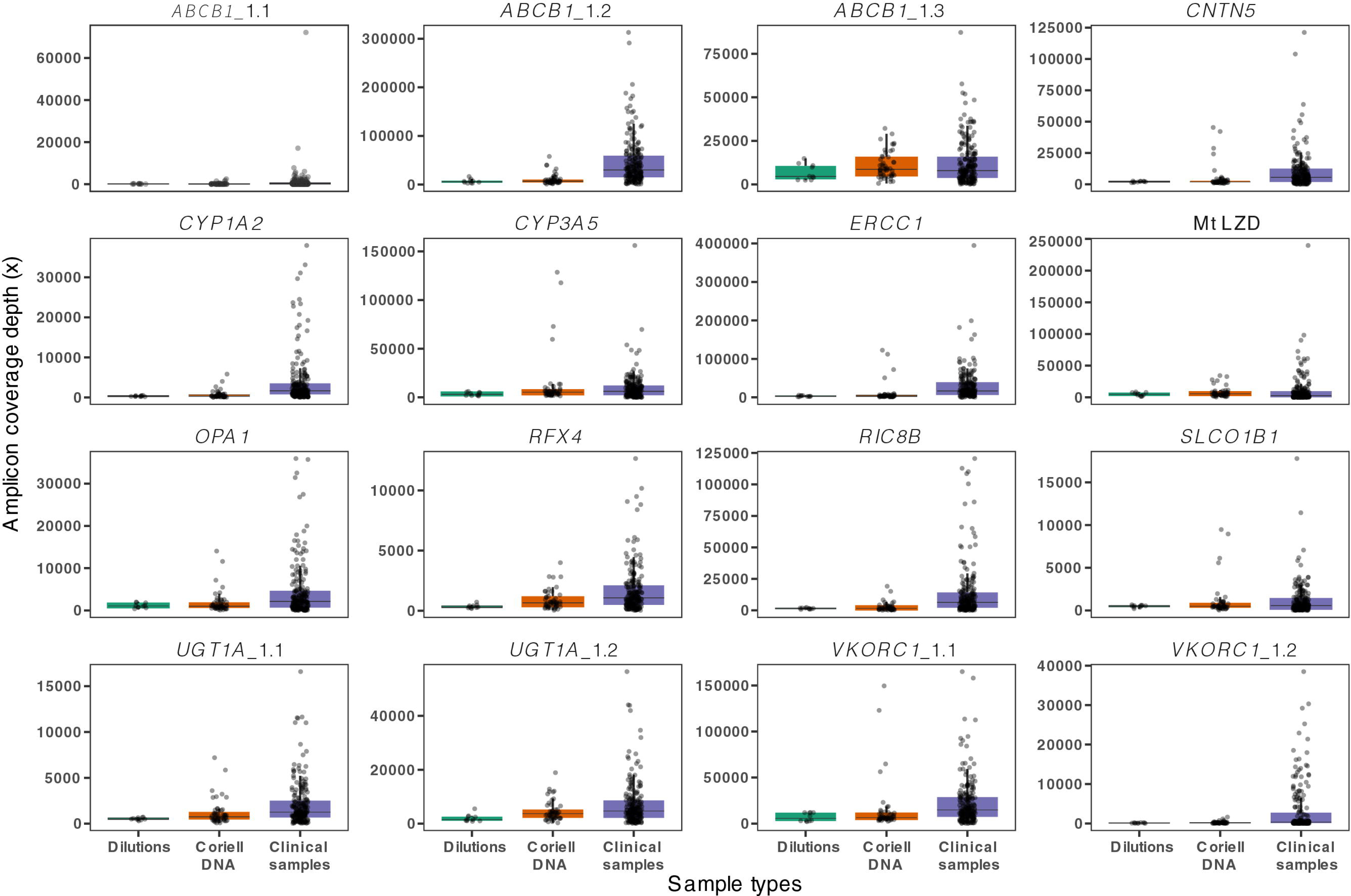
Gene coverage across different sample types using the targeted Nanopore pharmacogenomics sequencing panel. This panel targets 23 SNPs across 12 genes using 16 amplicons. The data set includes 5 dilutions (100, 50, 25, 12.5, and 6.25 ng/µL) prepared in replicates from a Coriell DNA sample sequenced on a Flongle flow cell and a SpotON flow cell, and other 50 Coriell DNA samples from the 1000 Genome project and 202 clinical samples. Each boxplot represents the coverage distribution for a specific amplicon across the different sample types.

### Clinical characteristics of the Indian MDR-TB cohort participants

Participants had a median age of 27 years (IQR: 21-35), with 18 individuals <18 years of age (8.9%), and a median weight of 55 kg (IQR: 46-65). Overall, 64.9% of participants were female. Most patients had pulmonary TB (64.3%), with cavitary lung disease identified among 41.6% and 25.7% reporting treatment for a previous episode of tuberculosis. Many participants were underweight, with BMI<18.5kg/m2 (36.6%), comorbid diabetes affected 16.3%, while HIV was uncommon (2.5%), as were self-reported use of alcohol (7.4%) or tobacco (3.0%). Despite complex drug resistance, 87.6% of participants completed treatment without evidence of relapse, while 5% died of any cause during participation, 4% either discontinued therapy or were lost to follow-up, 3% were transferred to other centers, and 0.5% experienced relapse. Clinical characteristics of study participants are presented in **Table 1**.

### Nanopore sequencing of saliva samples from clinical cohort

Sequencing depth exceeded the minimum cutoff (>50X) for all amplicons in 202 saliva samples. Median sequencing depth across the 16 amplicons was 3,609X (IQR: 1,207-7,578), with 94.8% of amplicons having depth greater than 100X and 81.0% greater than 500X **(Figure 2, Supplementary Table 3)**. The highest frequency of homozygous alternate alleles (189/202, 93.5%) was observed for the linezolid-associated mitochondrial variant A2706G, followed by *CYP3A5* rs776746 T>C (95/202, 47.0%) and *OPA1* rs1061648 A>G (80/202, 39.6%), which are associated with both linezolid and bedaquiline exposure and with ethambutol exposure, respectively (**Table 2**). The global distribution of the mutation frequencies of these variants across different populations is presented in **Figure 1B**.

**Table 2:**
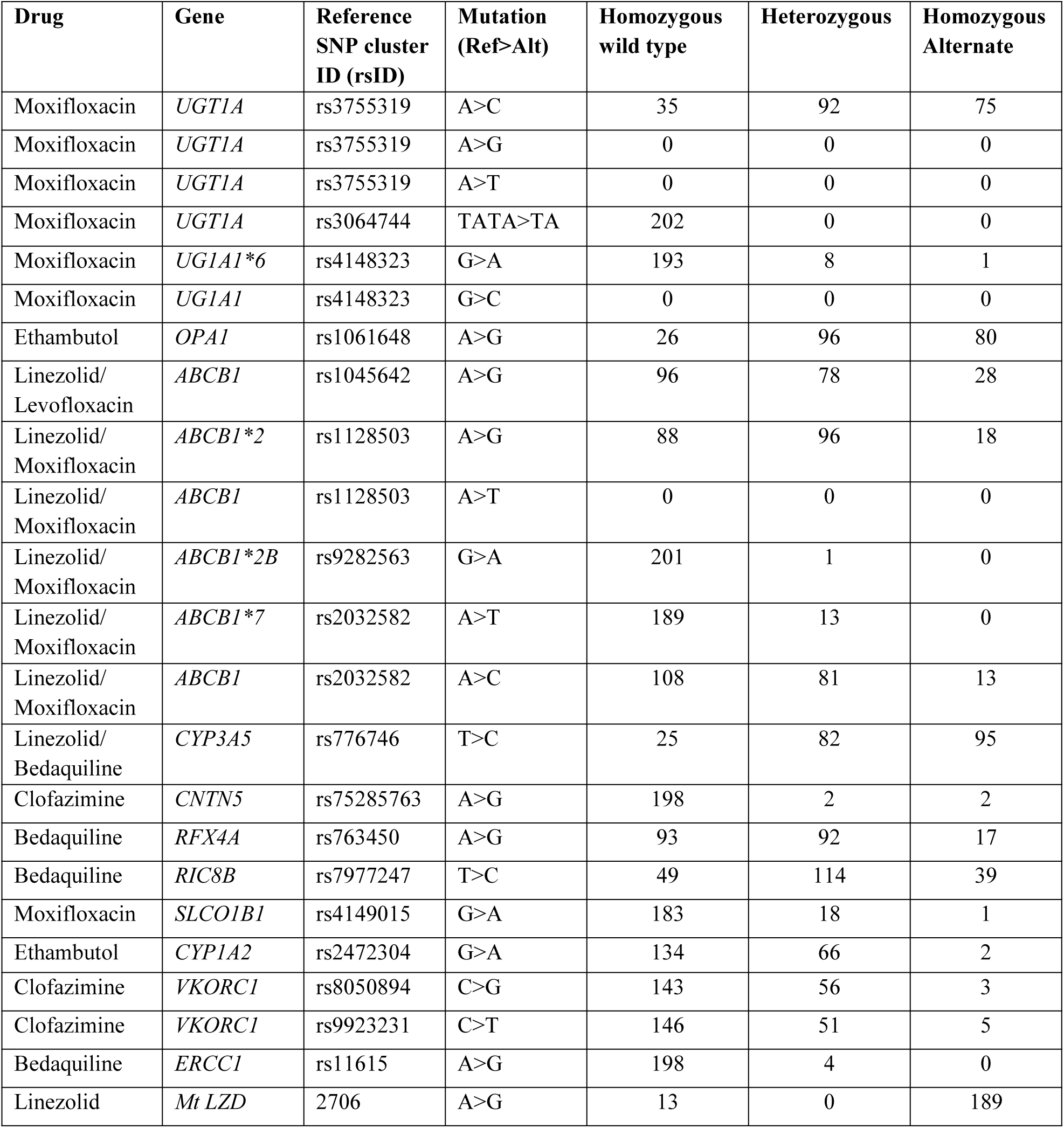
Distribution of genotypes across 23 single-nucleotide polymorphisms (SNPs) in clinical samples (n=202)

### Association of pharmacogenomic variants with drug concentrations

Of 174 drug-dose combinations tested against all 23 variants, 15 drug-dose combinations involving moxifloxacin and linezolid showed suggestive genotype-phenotype association trends with eight variants on the PGx panel **(Supplementary Figure 4)**. These trends were not seen with usual doses of linezolid or moxifloxacin (600mg and 400mg daily, respectively) or in the primary analysis including only the first measured steady-state Cmin concentration, but were found among subsets of individuals receiving different doses in the sensitivity analysis that included serial measurements throughout treatment for each participant. No significant associations were observed for the remaining drug-dose combinations and variants.

Among participants who experienced linezolid-associated toxicity while taking 600mg and were subsequently prescribed lower linezolid doses of 300 mg daily before their PK sampling (n=98), three *ABCB1* variants were associated with lower trough concentrations (Cmin). Homozygous alternate carriers at rs1045642 A>G (12/98) had significantly lower Cmin than wild-type carriers (50/98; p<0.05). For rs2032582 A>C, homozygous alternate carriers (7/98) differed significantly from both wild type (p<0.05) and heterozygous carriers (p<0.01), indicating reduced linezolid exposure in homozygous alternate individuals following dose reduction. For rs2032582 A>T, the rare homozygous alternate carrier (1/98) also differed significantly from wild type (p<0.05) and heterozygous carriers (p<0.01), again consistent with lower exposure in homozygous alternate individuals. At rs1128503 A>G, Cmin after dose reduction differed between heterozygous (41/98, p<0.05) and homozygous alternate carriers (11/98, p<0.05) participants, whereas comparisons involving wild-type carriers (46/98 participants) were not significant. At the 600 mg daily dose, no genotype-specific differences in Cmin reached statistical significance (all p≥0.05) (**Figure 3A**).

**Figure 3A:**
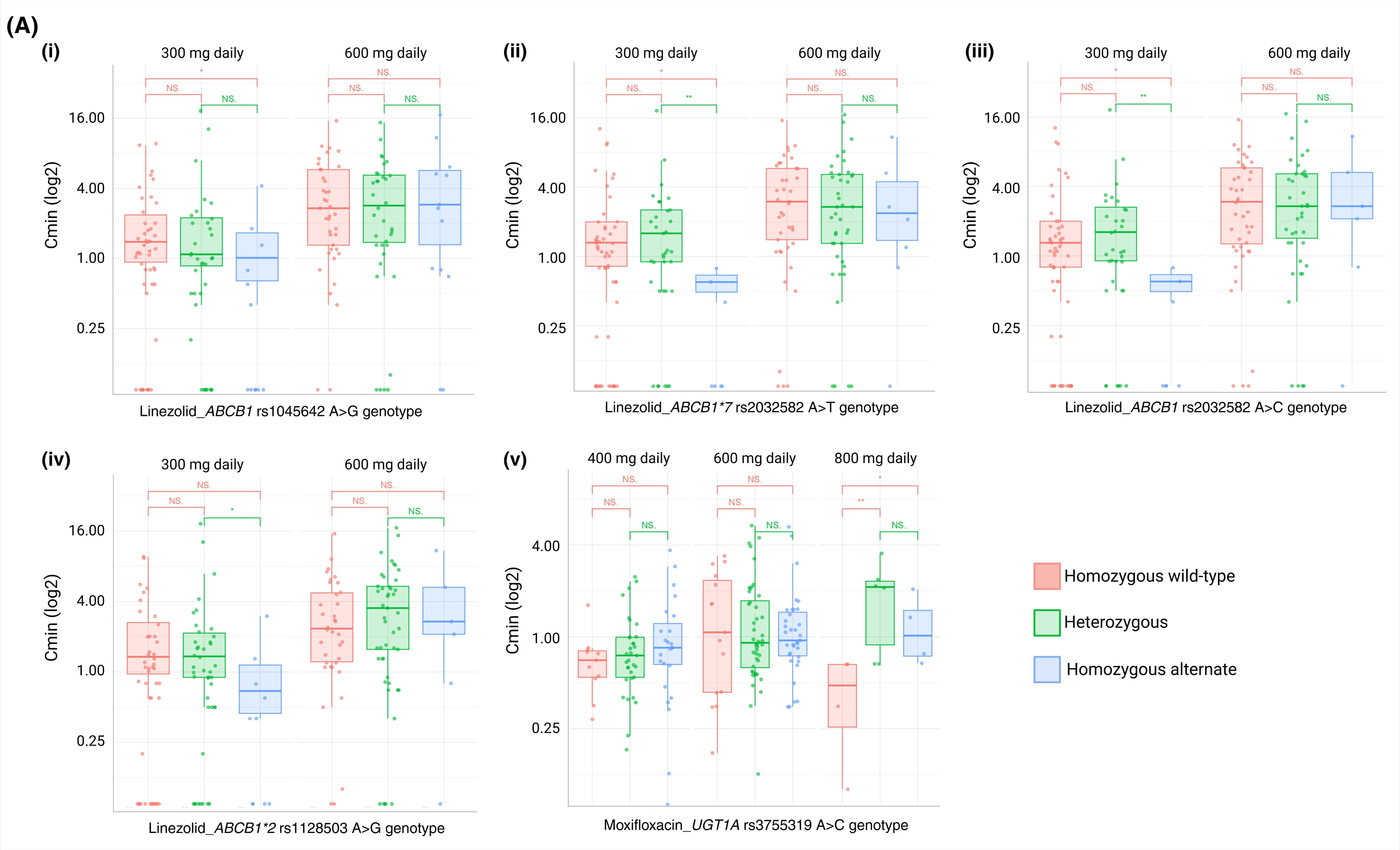
Box and whisker plots of first trough drug levels for each drug tested, stratified by pharmacogenomic variants.) (i) *ABCB1* rs1045642 A>G; (ii) *ABCB1*7* rs2032582 A>T; (iii) *ABCB1* rs2032582 A>C; (iv) *ABCB1*2* rs1128503 A>G; and (v) *UGT1A*6* rs3755319 A>C. Bars at the top indicate p-values from the Wilcoxon rank-sum test, with labels denoting NS (not significant), * (p < 0.05), or ** (p < 0.01).

After normalizing all participants prescribed linezolid to a 600mg daily equivalent exposure (doubling the measured concentrations of those receiving 300mg daily and analyzing them along with those receiving 600mg daily, n=187), significant differences in Cmin were seen between heterozygous and homozygous alternates for *ABCB1* rs2032582 A>T (174/10/3), rs2032582 A>C (96/77/14), and rs1128503 A>G (82/88/17) (all p<0.05), though, indicating a modest transporter-mediated reduction in exposure among homozygous alternates, though no difference was not seen between homozygous reference and heterozygous individuals in this analysis. Clinically, the observed effects are modest, limited to selected genotype contrasts, and derived from relatively few homozygous-alternate individuals, they therefore serve primarily as preliminary signals implicating *ABCB1* rs2032582 A>T/C and rs1128503 A>G as priorities for future PK-PD and outcome studies, rather than as evidence to support immediate genotype-guided dose adjustment (**Figure 3B**).

**Figure 3B.**
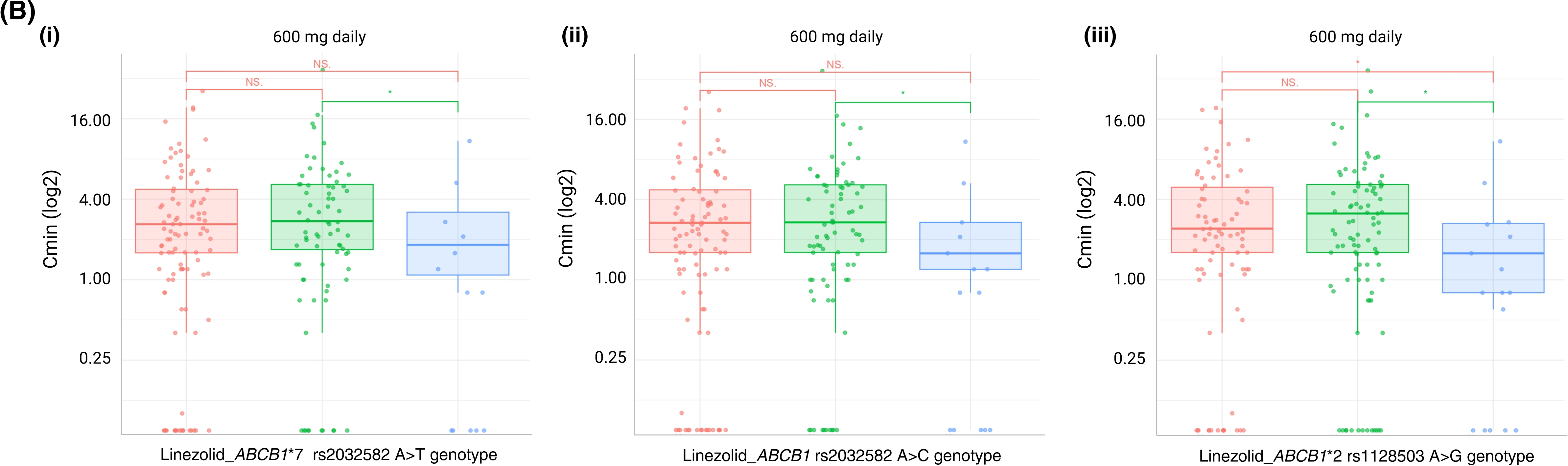
Secondary analysis of linezolid exposure after dose normalization to 600 mg (i) *ABCB1*7* rs2032582 A>T; (ii) *ABCB1* rs2032582 A>C; and (iii) *ABCB1*2* rs1128503 A>G. Dots overlying each box plot represent individual values. Bars at the top indicate p-values from the Wilcoxon rank-sum test, with labels denoting NS (not significant), * (p < 0.05), or ** (p < 0.01).

For moxifloxacin, *UGT1A1* rs3755319 A>C did not show significant genotype effects at standard doses. At 400mg and 600mg, trough concentrations were comparable across genotypes, with no pairwise comparisons reaching significant differences. In contrast, at 800mg (n=14: 4 wild-type, 6 heterozygous, 4 homozygous alternate), trough concentrations were significantly higher in variant carriers, with differences between wild-type and heterozygous (p<0.01) and between wild-type and homozygous alternate (p<0.05). These findings suggest a possible dose-related effect of *UGT1A1* rs3755319 A>C on moxifloxacin exposure at the extremes of dosing, however, given the small sample size and lack of a consistent signal at standard doses, these results require confirmation in larger cohorts and dose-normalized population PK analyses before any clinical implications are inferred (**Figure 3A**).

### Pharmacogenomic determinants of anti-TB drug toxicity

Of the 152 adverse drug reaction combinations analyzed, eight toxicity profiles associated with linezolid, moxifloxacin, and clofazimine showed significant trends (**Supplementary Table 5**). Although previous studies have reported associations between certain variants and adverse drug reactions, the majority of variants screened in our study did not show significant genotype-toxicity associations. Among the individuals in whom statistically significant genotype–toxicity associations were observed, several genotype categories were represented by only a small number of participants despite reaching statistical significance, preventing assessments among subgroups and dose-specific toxicity.

Linezolid toxicity analyses revealed two main genotype-toxicity signals. The *ABCB1* rs1045642 A>G polymorphism was associated with a higher frequency of linezolid-associated toxicity (any grade) during prolonged therapy. Toxicity was observed in 36.1% (26/72) of heterozygous and 42.3% (11/26) of homozygous alternate participants, compared with 23.6% (21/89) of participants with the homozygous reference genotype (p=0.03). Similarly, carriers of the ABCB1 rs2032582 A>T variant showed a higher frequency of any linezolid-associated toxicity (58.3%, 7/12) than homozygous reference carriers (29.1%, 51/175; p=0.03).

Among patients treated with moxifloxacin, the *ABCB1*7* rs2032582 A>T variant was associated with joint pain at any severity, with frequencies of 55.6% (5/9) among heterozygous and 100% (2/2) among homozygous alternate carriers, compared with 36.4% (60/165) among homozygous reference carriers (p=0.04). This variant was not associated with high-grade (3+) joint pain (p=0.73). In addition, the *UGT1A1*6* rs4148323 G>A variant was associated with the frequency of any moxifloxacin-associated toxicity, occurring in 42.9% (3/7) of heterozygous individuals compared with 14.3% (24/168) of homozygous reference carriers (p=0.04), though the variant was significantly associated with a higher frequency of transaminase elevation, QTc prolongation, or joint pain of any grade when measured independently.

Among the variants analyzed, *CYP3A5* rs776746 T>C previously reported to influence linezolid-associated toxicity showed fair representation across genotype groups (homozygous reference: 7/17, 41.2%; heterozygous: 29/82, 35.4%; homozygous alternate: 22/88, 25%), but no significant difference in toxicity was observed in our cohort (p=0.0857).

## DISCUSSION

Pharmacogenomic screening of patients for subtherapeutic trough concentrations and treatment-associated toxicities of anti-TB drugs prior to treatment initiation could enable personalized dosing and improve outcomes in some patients. However, pharmacogenomic testing has not yet been integrated into routine TB care mainly due to a lack of pharmacogenomic screening tests and limited evidence of clinically actionable findings beyond testing for NAT2 genotypes. To address this gap, we developed a scalable, cost-effective pharmacogenomic sequencing panel on a portable Oxford Nanopore platform to detect host variants associated with subtherapeutic exposure and toxicity for second-line drugs. We first demonstrated 100% concordance with Illumina whole-genome sequencing, then clinically validated the panel on saliva samples from 202 cohort participants treated for active MDR-TB.

Although most variants were not associated with large shifts in exposure across all doses, Nanopore-derived genotypes showed suggestive genotype-exposure trends for moxifloxacin and linezolid, two key agents within the BPaLM regimen. *ABCB1* polymorphisms were associated with lower linezolid trough concentrations at 300mg, and *UGT1A* rs3755319 A>C was associated with moxifloxacin exposure at 800mg. These findings suggest that pharmacogenomic information, combined with therapeutic drug monitoring, could be useful for targeted safety surveillance or dose optimization in defined high-risk groups. However, the observed effects are limited to selected genotype contrasts and derived from relatively few homozygous-alternate individuals, indicating that these signals are promising but require targeted validation before informing dosing practice.

PGx-guided dosing is increasingly recognized as a strategy to optimize TB therapy, but most work has focused on first-line drugs and has revealed substantial ethnic differences in variant frequencies and drug metabolism (34, 35). For example, the functional *NAT2* 191G>A variant, common in many African populations, is omitted from existing prediction tools such as NAT2Pred, leading to misclassification of isoniazid acetylator status (34). This example illustrates how assays developed in limited ancestry groups may perform poorly when generalized. Similarly, Chigutsa et al. showed that *SLCO1B1* rs4149032 G>A, which is highly prevalent among people with tuberculosis in South Africa, was associated with an 18-28% reduction in rifampin bioavailability (36). In contrast, Swaminathan et al. reported that rs4149032 G>A in *SLCO1B1* did not influence rifampicin plasma concentrations in adults from a South Indian cohort (37). These findings suggest that lack of association in one population does not preclude a clinically relevant effect in another and underscore the need for population-specific pharmacogenomic evaluation. In our study, we designed a PGx panel using markers reported across diverse populations to minimize population-specific bias and provide the first description of TB pharmacogenomic diversity in an Indian cohort relative to global data.

We found that although moxifloxacin exposure was not significantly associated with *UGT1A* rs3755319 A>C at standard doses of 400 or 600 mg daily, among participants prescribed 800 mg daily, trough concentrations were lower in wild-type carriers than in heterozygous or homozygous-alternate carriers. The *UGT1A1* gene encodes a key glucuronidation enzyme, and its reduced activity can lead to slow drug clearance. Studies have reported that *UGT1A1* rs3755319 A>C can reduce *UGT1A1* expression or activity, with wild-type or heterozygous individuals having approximately 11% higher clearance than homozygous alternate carriers (5, 38).

Another important gene that was found to be significant in our study was *ABCB1*. The *ABCB1* gene encodes the drug transporter P-glycoprotein, an efflux pump that regulates absorption, distribution, and intracellular concentrations of anti-TB drugs. Several studies have reported the impact of *ABCB1* variants on linezolid pharmacokinetics (39, 40). Sarah et al. showed that the *ABCB1* c.3435C>T (rs1045642 A>G) variant significantly reduced linezolid clearance (13.19 vs 7.82 L/h for CC vs CT/TT), with corresponding increases in half-life (2.78 vs 5.45 h) and volume of distribution (37.43 vs 46.71 L) (41). Another study found that while variant *ABCB1* rs1128503 A>G does not appear to alter P-glycoprotein structure, it may affect substrate specificity and pharmacokinetics (42). In this study, we observed that participants carrying variants in *ABCB1* whose doses had been reduced to 300mg in the setting of linezolid-associated toxicity had significantly lower trough concentrations of linezolid. This may reflect an impact of the variant that increases efflux to a degree that is not clinically significant at normal dosing (600mg daily), but for which heterozygous alternate status may result in lower than desired doses for those dose-reduced to 300mg daily. In our study, both homozygous alternate and heterozygous carriers of *ABCB1* rs1128503 A>G exhibited lower Cmin at a 300mg daily dose compared with wild-type carriers. These findings suggest that in patients who require dose reduction to 300mg, *ABCB1* genotype may identify subgroups at risk of underexposure, and future studies should explore pharmacogenetic-guided dosing or enhanced TDM in high-prevalence populations. However, in our analysis, these differences remained within the therapeutic range for linezolid (2-8 mg/L) in patients receiving 600mg daily, suggesting only minimal impact on exposure. Among the predicted *ABCB1* variants, which encode the efflux transporter P-glycoprotein, we provide the first clinical evidence that the *ABCB1*7* rs2032582 A>T variant affects linezolid plasma trough concentrations for those whose doses have been reduced due to pre-existing toxicity. To our knowledge, this is the first pharmacogenomic analysis linking second-line anti-TB drug exposure to the predicted *ABCB1*7* rs2032582 A>T variant that can be validated in larger studies.

For the majority of markers tested in the toxicity analysis, we did not observe significant clinical associations. One possible explanation is that, unlike drug-susceptible TB treatment, the high frequency of dose combinations and adjustments in MDR-TB treatment requires large numbers of patients to evaluate clinical associations between variants and toxicity at each drug exposure. Our cohort lacked adequate representation of variants at several genomic locations, thereby limiting statistical power. We did observe nominal significance for a few host genetic variants in relation to linezolid-and moxifloxacin-induced toxicity, although the numbers remained modest. Variants in the *ABCB1* gene were associated with linezolid-induced toxicity of any grade, consistent with previous reports. For instance, prior studies have shown that linezolid is associated with tissue-specific toxicity, including nerve fiber swelling, hematologic abnormalities, and peripheral neuropathy (43). Earlier studies have also reported lactic acidosis with long-term linezolid exposure among individuals carrying the A2706G mitochondrial DNA polymorphism (44, 45). However, we did not observe significant differences for the mitochondrial marker tested in our cohort. Larger cohort studies are needed to confirm these findings. For moxifloxacin, we observed an association between *UGT1A1* genetic variants and overall drug-induced toxicity. While this was not driven by any individual toxicity we monitored, these results are consistent with previous studies showing that *UGT1A1* variants reduce moxifloxacin clearance and may increase the risk of liver injury (5, 38), though this trend.

Leveraging genotype information to predict plasma drug levels and toxicities has been demonstrated for isoniazid but largely lacks for other drugs (35). Most existing pharmacogenomic assays, however, use qPCR. We previously developed a GeneXpert cartridge-based assay using five SNPs identified from random-forest models that predicted isoniazid metabolism with high accuracy (34). Although rapid and user-friendly, PCR-based assays accommodate only a small number of variants and generate unphased data. Given that TB treatment involves multiple drugs and numerous PGx markers that vary across populations, broader and more flexible approaches are needed. Targeted next-generation sequencing directly addresses these limitations. A targeted panel can screen many variants simultaneously and can be easily updated as new markers emerge, without major additional cost. In 2024, WHO endorsed targeted sequencing for drug-resistant TB, prompting global investments in sequencing infrastructure and workforce training (46). This expanding capacity provides a natural entry point for implementing Nanopore-based PGx panels within existing TB diagnostic workflows.

Our group previously developed a targeted sequencing panel for rifampicin and isoniazid and evaluated it against pharmacokinetic data from a South African clinical cohort (35). Variants captured by this panel significantly predicted plasma levels of both drugs, supporting the role of tNGS in dose individualization. Nanopore-based tNGS offers additional advantages: relatively low capital costs (approximately 3,000 USD for the sequencer), availability of small and inexpensive flow cells that allow sequencing in small batches, and real-time run monitoring, so sequencing can be stopped once sufficient coverage is reached. In this study, we compared the larger SpotON flow cell with the smaller Flongle and showed that the required coverage could be achieved on the smaller flow cell without compromising sequencing quality, further supporting the feasibility of Nanopore-based tNGS for pharmacogenomic implementation in TB programs.

This study has several limitations. First, because this was a population genomic study in which the frequencies of many pharmacogene variants were previously unknown in India, the cohort was underpowered for robust assessments of genotype-drug level associations. Now that variant frequencies have been defined in this setting, larger studies are needed to validate their impact on common toxicities at the population level. Second, MDR-TB treatments are highly individualized both in component drugs and doses, which makes it difficult to isolate pharmacogenomic effects from other determinants of exposure. Evaluation of these effects with appropriate adjustments for multiple comparisons needs to be assessed through larger, adequately powered studies in the future. Additionally, the pharmacogenomic associations presented here relied on pre-dose (trough) concentrations without direct observation of the previous dose rather than full pharmacokinetic profiles. Single-timepoint sampling carries substantial uncertainty, and dose normalization only coarsely adjusts for dosing differences, failing to capture nonlinear dose-clearance-exposure relationships or covariates like concomitant drug interactions, renal function, and body weight. Properly addressing this requires drug-specific population pharmacokinetic modelling incorporating full concentration-time profiles and relevant covariates, which was beyond this study’s scope and warrants future population pharmacokinetic confirmation. The Indian subcontinent comprises multiple, genetically distinct populations, and our cosmopolitan cohort included individuals from several ethnic groups; further work in more homogeneous subpopulations may clarify the distribution and impact of these pharmacogenes across India. Finally, we restricted the panel to pharmacogenes with matched plasma drug level data from available clinical samples, so although the assay is expandable, it does not yet capture all second-line drugs used in MDR TB regimens.

## CONCLUSIONS

Our study demonstrates that targeted Nanopore sequencing is a reliable and effective approach for detecting pharmacogenomic polymorphisms associated with the pharmacokinetics of anti-TB drugs. The pharmacogenomic panel developed here showed complete concordance with Illumina sequencing and achieved high coverage in clinical samples, highlighting its accuracy and feasibility for real-world application. This approach highlights the potential of Nanopore sequencing to enable rapid, low-cost, and portable genotyping oriented around promising pharmacogenes that may support personalized dosing.

## Data availability

The data that support the findings of this study are available on request from the corresponding author.

## Author contributions

R.V., T.A., J.A.T., and A.P. conceptualized and designed the study. P.Y. and R.V. developed the panel. P.Y., R.V., S.S., A.S.B., and M.P. performed the experiments. A.S.B. and S.V. prepared the figures and tables. K.V. and J.A.T. performed the data analysis. P.R.A., R.V.L., and H.U.B.P. recruited patients and collected data. R.V. and P.Y. wrote the first draft. C.R., P.D., J.R.A., A.P., and T.A. provided input on data analysis and interpretation. All authors reviewed the final manuscript, provided feedback, and approved the final version.

## Data Availability

All data produced in the present study are available upon reasonable request to the authors

## Acknowledgements

The authors acknowledge the technical staff and patients from the clinical site for their cooperation during the study. The authors would like to acknowledge Ivan Nkuhairwe for his help with the data handling.

The MDR-TB MUKT Study Team: Lancelot M. Pinto, Jai B. Mullerpattan, Ayesha Sunavala, Amita Gupta, Jigneshkumar Patel, Alpa J. Dherai, Bhamini Keny, Suvarna Gaikwad, Namrata Sawant.

---

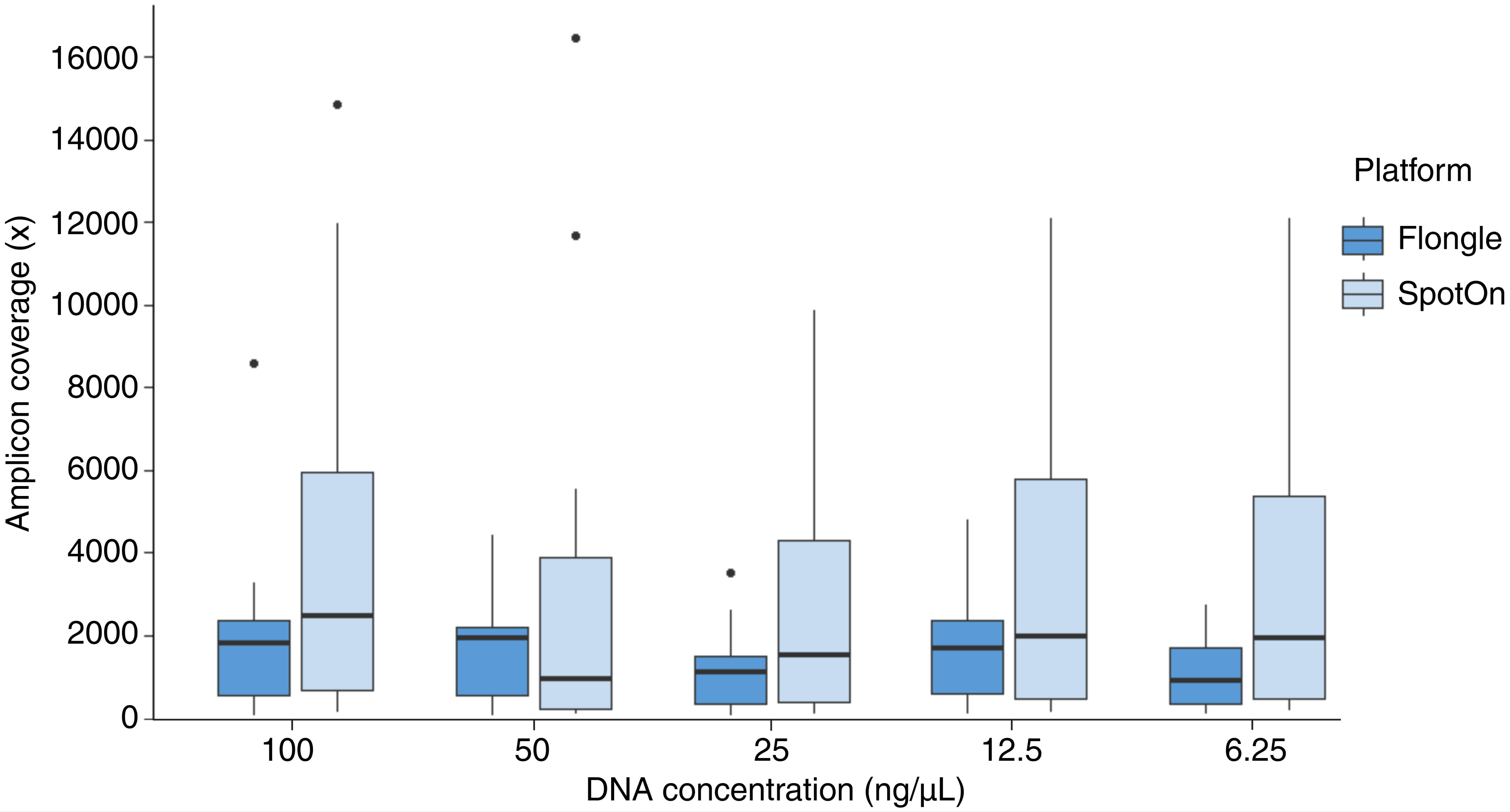

