## Supplementary material for "Nanopore sequencing panel for saliva-based host pharmacogenomic screening in anti-tubercular therapy": Online supplement

**Targeted Nanopore sequencing for saliva-based screening of host pharmacogenes for anti-TB drugs**

**Online supplement**

**METHODS**

**Primer design**

To perform targeted sequencing, we used a multiplex strategy that relied on anchored primers. This involves amplifying the regions of interest using specific primers flanked by Nanopore-compatible anchor sequences (5’-Anchor- TTTCTGTTGGTGCTGATATTGC and 3’-Anchor- ACTTGCCTGTCGCTCTATCTTC). The anchor sequences enable a second round of PCR with Oxford Nanopore's barcoded outer primers for rapid adapter attachment. The second PCR uses fewer cycles and creates barcoded amplicons with modified 5' ends for simplified post-PCR adapter attachment. We developed a 16-plex panel amplifying regions in 12 genes. Primers were designed to amplify products between 445bp-826bp using Beacon Designer (Premier Biosoft International, version 8.21). Primer specificity was verified with Primer-blast (NCBI). Primer concentrations and sequence details are listed in **supplementary table 2.**

**Monitoring adverse drug reactions**

Scheduled clinical evaluations were performed following treatment initiation at monthly intervals for the first six months, then quarterly until the end of therapy. At each evaluation, participants were assessed for multiple toxicities, including QT interval prolongation assessed by ECG twice-yearly and measured by Fridericia criteria, neuropathy by self-report as well as 5g monofilament and 128Hz vibration testing of both feet, visual acuity by Snellen and Ishihara plates, self-reported change in skin color, and by scheduled monitoring of transaminases and complete blood cell counts. Each toxicity was classified as “ever occurring during therapy” and as “ever occurring during therapy at a grade of 3 or higher” following DAIDS categorization.

We analyzed the associations between pharmacogenetic markers and treatment-associated toxicity. Each toxicity is listed by drug and, where applicable, categorized as either "any" or "Grade 3+." For example, linezolid is associated with bone marrow suppression, which can manifest as anemia, thrombocytopenia, and/or leukopenia. We categorized all individuals taking linezolid based on whether they developed a reduction in any blood cell lines below normal ranges (“any cytopenias”) and whether they experienced Grade 3 or higher anemia, thrombocytopenia, or leukopenia (“cytopenia [Grade 3+]”), according to results for each linezolid-associated pharmacogenetic marker. In several cases, apparent differences between pharmacogenetic groups could not be statistically analyzed using the Kruskal-Wallis test due to the absence of participants with a particular genotype and toxicity combination.

**DNA extraction from saliva samples**

Saliva samples (n=202) stored at -80°C from the MUKT cohort of Hinduja Hospital, Mumbai were used for the validation of the panel on clinical samples. The DNA was extracted using QIAamp DNA Blood Mini Kit **(**Cat # 51304) according to the manufacturer’s instructions. Samples were gradually thawed from -80ºC to -20ºC to room temperature before DNA extraction. Briefly, 1mL of saliva sample was mixed with 4mL of PBS (1:4 ratio) and centrifuged at 4000 RPM for 5 minutes. The supernatant was discarded, and the pellet was resuspended in 180μL of PBS. Then, 20μL of QIAGEN Protease and 200μL of Buffer AL were added, and the sample was immediately vortexed for 15 seconds to ensure efficient lysis. The mixture was incubated at 56°C for 20 minutes. Afterwards, 200μL of ethanol (96-100%) was added, and the sample was vortexed again. Following a brief centrifugation to remove droplets from the lid, the mixture was applied to a QIAamp Spin Column placed in a 2mL collection tube and centrifuged at 8000 RPM for 1 minute. The column was then washed with 500μL of Buffer AW1, followed by 500μL of Buffer AW2, with centrifugation after each wash. A final spin at full speed was performed to ensure complete removal of Buffer AW2. The column was transferred to a clean 1.5mL tube, and DNA was eluted with 50μL of nuclease-free water. After a 10-minute incubation at room temperature, a final centrifugation was carried out, and the eluted DNA was stored at -20°C till further use.

**MinION library preparation and sequencing**

For panel development and validation, we sequenced a total of 50 Coriell DNA samples plus one Coriell DNA sample at 5 dilutions (100ng/μL, 50ng/μL, 25ng/μL, 12.5ng/μL, and 6.25ng/μL) using both Flongle and SpotON flow cells. For clinical pharmacogenomic validation, we used 202 frozen saliva samples from the MUKT cohort of PD Hinduja Hospital. We used two types of flow cells, a smaller MinION Flongle flow cell with up to 2.8Gb output, and a regular SpotON R10.4.1 flow cell with 20Gb output. The sequencing statistics of the runs are provided in **Supplementary Table** **3**. The library was prepared using the SQK-LSK114 Ligation Sequencing Kit (Oxford Nanopore Technologies). Samples were barcoded using a Nanopore PCR barcoding expansion (EXP-PBC096 PCR Barcoding Expansion). For each library, we took approximately 100ng of the purified PCR product from the first round of PCR for barcoding. The barcoding mix was prepared by adding 1μL of 10uM PCR barcode to 25 μL LongAmp Taq 2x master mix (NEB) and 100ng of first-round PCR product. The volume was adjusted to 50μL with nuclease-free water, and barcoding was performed at 12 rounds of PCR cycles. The purified libraries were pooled to a total 1μg & 1.5μg for Flongle and SpotON flow cells, respectively, followed by DNA repair and end-prep using NEBNext FFPE DNA Repair Mix and NEBNext Ultra II End repair/dA-tailing Module reagents in accordance with the manufacturer’s instructions. Adaptor ligation was performed using Adapter Mix F (AMX-F) and Quick T4 Ligase. For targeted sequencing, we used 200μL Short Fragment Buffer (SFB) for the final wash to retain shorter amplicons. The final library was eluted in 15μL elution buffer (EB). We loaded the maximum recommended quantities of libraries in all runs (20fm libraries on a Flongle flow cell and 75fm of a SpotON flow cell). The samples were sequenced on a MinION Mk1C sequencer.

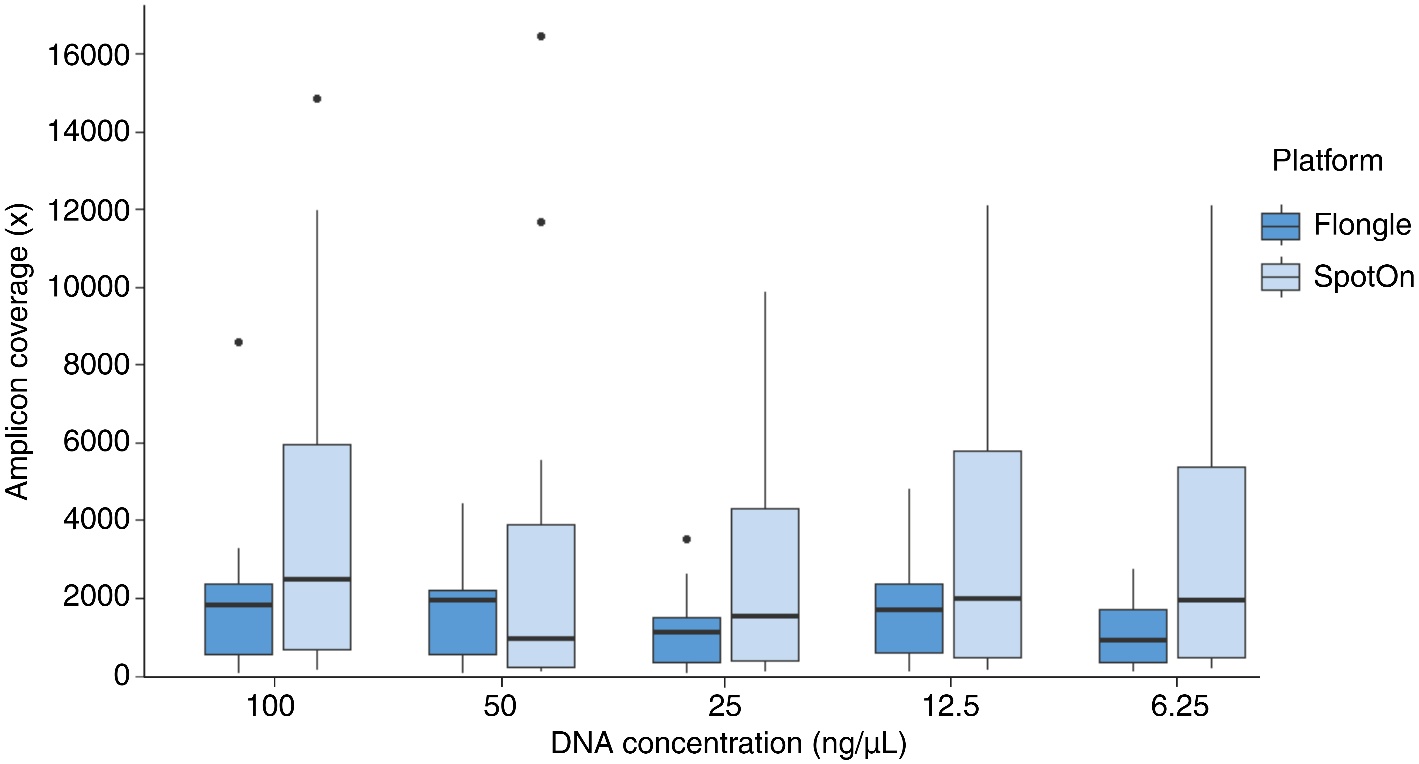

**Supplementary figure 1:** Amplicon coverage across different dilutions (100, 50, 25, 12.5, and 6.25ng/µL) prepared in replicates from a Coriell DNA sample sequenced on the Flongle flow cell and SpotON flow cell.
