## Supplementary Table 2 for "Nanopore sequencing panel for saliva-based host pharmacogenomic screening in anti-tubercular therapy"

Table S2: List of anchored primers used (n=16) in this study

| **Antibiotics** | **Gene** | **SNP position** | **Primers** | **Product Length** | **Tm** | **Anchored Sequence** |
| --- | --- | --- | --- | --- | --- | --- |
| Linezolid/  Moxifloxacin | *ABCB1* | rs2032582 | ABCB1_1.1_FP | 567 | 63 | TTTCTGTTGGTGCTGATATTGCGAAGGAAGAACAGTGTGAAGA |
|  |  |  | ABCB1_1.1_RP |  | 63 | ACTTGCCTGTCGCTCTATCTTCTAGAAGCATGAGTTGTGAAGATAA |
| Linezolid/  Levofloxacin | *ABCB1* | rs1045642 | ABCB1_1.2_FP | 601 | 63.9 | TTTCTGTTGGTGCTGATATTGCGTGGAGCCTCAAGCCTATA |
|  |  |  | ABCB1_1.2_RP |  | 63.6 | ACTTGCCTGTCGCTCTATCTTCGTGCTGGTCCTGAAGTTG |
| Linezolid/  Moxifloxacin | *ABCB1* | rs1128503 | ABCB1_1.3_FP | 826 | 62.5 | TTTCTGTTGGTGCTGATATTGCTATCGTGGTGGCAAACAA |
|  |  |  | ABCB1_1.3_RP |  | 63.2 | ACTTGCCTGTCGCTCTATCTTCAATTGATAATGTAAGTCTGAGTTGG |
| Linezolid/  Bedaquiline | *CYP3A5* | rs776746 | CYP3A5_FP | 639 | 62.8 | TTTCTGTTGGTGCTGATATTGCGGGAGTTGACCTTCATACG |
|  |  |  | CYP3A5_RP |  | 63.2 | ACTTGCCTGTCGCTCTATCTTCCAAGTCCTCAGAATCCACAG |
| Bedaquiline | *RFX4* | rs763450 | RFX4_FP | 663 | 63.2 | TTTCTGTTGGTGCTGATATTGCTGAAACATGCTCTGCCTTT |
|  |  |  | RFX4_RP |  | 63.1 | ACTTGCCTGTCGCTCTATCTTCCCTCTTGGTTACATTTGACTTATTC |
| Bedaquiline | *RIC8B* | rs7977247 | RIC8B_FP | 476 | 62.1 | TTTCTGTTGGTGCTGATATTGCCATACATCTGTCAGGTAGTTCT |
|  |  |  | RIC8B_RP |  | 62.2 | ACTTGCCTGTCGCTCTATCTTCAAGACTTGAAGGTGGTGAG |
| Bedaquiline | *ERCC1* | rs11615 | ERCC1_FP | 445 | 63.5 | TTTCTGTTGGTGCTGATATTGCAGAGGCTTCTCATAGAACAGT |
|  |  |  | ERCC1_RP |  | 63.2 | ACTTGCCTGTCGCTCTATCTTCCAGGGTTAGGAGGAGAGAG |
| Moxifloxacin | *SLCO1B1* | rs4149015 | SLCO1B1_FP | 546 | 62.5 | TTTCTGTTGGTGCTGATATTGCTGGGTCTACATTTCTCAGTTC |
|  |  |  | SLCO1B1_RP |  | 62.5 | ACTTGCCTGTCGCTCTATCTTCTTATTGACTTGACTTGTGGAGA |
| Moxifloxacin | *UGT1A1* | rs3755319 | UGT1A_1.1_FP | 645 | 64 | TTTCTGTTGGTGCTGATATTGCCAAAACTCTAGTTAGCTGTTTCTCT |
|  |  |  | UGT1A_1.1_RP |  | 63.3 | ACTTGCCTGTCGCTCTATCTTCGCTTCATCAACCAATCAGAATG |
| Moxifloxacin | *UGT1A1* | rs3064744, rs4148323 | UGT1A_1.2_FP | 757 | 64.2 | TTTCTGTTGGTGCTGATATTGCTGCTGTGTTCACTCAAGAATG |
|  |  |  | UGT1A_1.2_RP |  | 64.3 | ACTTGCCTGTCGCTCTATCTTCAAGGAAAGGGTCCGTCAG |
| Clofazimine | *CNTN5* | rs75285763 | CNTN5_FP | 526 | 63.9 | TTTCTGTTGGTGCTGATATTGCACACAGCCAAATCATATTATTCCA |
|  |  |  | CNTN5_RP |  | 64.4 | ACTTGCCTGTCGCTCTATCTTCCAAACAATGCCAGCCCAT |
| Clofazimine | *VKORC1* | rs8050894 | VKORC1_1.1_FP | 732 | 64.3 | TTTCTGTTGGTGCTGATATTGCGGCTAAGGTGGGAGGATC |
|  |  |  | VKORC1_1.1_RP |  | 63.5 | ACTTGCCTGTCGCTCTATCTTCGGGAGGATAGGGTCAGTG |
| Clofazimine | *VKORC1* | rs9923231 | VKORC1_1.2_FP | 786 | 63.1 | TTTCTGTTGGTGCTGATATTGCTCAAGTTTTTGGAAAGATTCTACAC |
|  |  |  | VKORC1_1.2_RP |  | 63.3 | ACTTGCCTGTCGCTCTATCTTCAGGAGATTGGTCAGCTTAATTC |
| Ethambutol | *CYP1A2* | rs2472304 | CYP1A2_FP | 563 | 63.6 | TTTCTGTTGGTGCTGATATTGCAGCAGATACTTGGGAAATGATG |
|  |  |  | CYP1A2_RP |  | 64.8 | ACTTGCCTGTCGCTCTATCTTCGAAGGTCTCCAGGATGAAGG |
| Ethambutol | *OPA1* | rs1061648 | OPA1_FP | 544 | 63.2 | TTTCTGTTGGTGCTGATATTGCCCTCTCGAATGGAAGAACAG |
|  |  |  | OPA1_RP |  | 62.9 | ACTTGCCTGTCGCTCTATCTTCATAGGTAAGCTGGTACTGACT |
| Linezolid | Mitochondrial genome | 2706 | MtLNZ_FP | 842 | 62.6 | TTTCTGTTGGTGCTGATATTGCAAGCAGCCACCAATTAAGA |
|  |  |  | MtLNZ_RP |  | 62.5 | ACTTGCCTGTCGCTCTATCTTCACCCTATTGTTGATATGGACTC |
