## Supplementary Table 3 for "Nanopore sequencing panel for saliva-based host pharmacogenomic screening in anti-tubercular therapy"

Table S3: Sequencing statistics for 262 (Coriell DNA, Dilutions, DNA from saliva) samples analyzed on Flongle and Spot on flow cell (R10.4.1)

| **Batch** | **Sample type** | **Total (n)** | **Amount of library (ng)/sample** | **Flow cell type** | **Run time (Hr.)** | **Active pores (n)** | **Median yield (Mb)/sample** | **Median depth (X)** |
| --- | --- | --- | --- | --- | --- | --- | --- | --- |
| Batch 1 | Coriell sample dilutions | 10 | 27 | Flongle | 24 | 77 | 30.0 | 1124 |
| Batch 2 | Coriell samples + dilutions | 55 | 25 | Spot On | 48 | 1200 | 69.3 | 1727 |
| Batch 3 | Saliva sample | 51 | 30 | Spot On | 72 | 1448 | 93.4 | 2257 |
| Batch 4 | Saliva sample | 10 | 100 | Flongle | 24 | 80 | 49.0 | 1234 |
| Batch 5 | Saliva sample | 54 | 28 | Spot On | 72 | 1555 | 182.0 | 8055 |
| Batch 6 | Saliva sample | 41 | 37 | Spot On | 72 | 1501 | 181.4 | 1562 |
| Batch 7 | Saliva sample | 44 | 34 | Spot On | 72 | 1591 | 312.5 | 11083 |
| Batch 8 | Saliva sample (Repeat) | 39 | 28 | Flongle | 24 | 81 | 16.1 | 403 |
| Batch 9 | Saliva sample | 6 | 26 | Flongle | 24 | 75 | 95.5 | 2044 |
